# Unresolved alterations in bile acid composition and dyslipidemia in maternal and cord blood after ursodeoxycholic acid treatment for intrahepatic cholestasis of pregnancy

**DOI:** 10.1101/2024.08.21.24312246

**Authors:** Srijani Basu, Sarah G. Običan, Enrico Bertaggia, Hannah Staab, M. Concepcion Izquierdo, Cynthia Gyamfi-Bannerman, Rebecca A. Haeusler

## Abstract

Intrahepatic cholestasis of pregnancy (ICP) is characterized by elevated plasma bile acid levels. ICP is linked to adverse metabolic outcomes, including a reported increased risk of gestational diabetes. The standard therapeutic approach for managing ICP is treatment with ursodeoxycholic acid (UDCA) and induction of labor prior to 40 weeks of gestation. To investigate bile acid and metabolic parameters after UDCA treatment, we enrolled 12 ICP patients with singleton pregnancies–half with and half without gestational diabetes–and 7 controls. Our study reveals that after UDCA treatment, notwithstanding a reduction in total bile acid and ALT levels, imbalances persist in the cholic acid (CA) to chenodeoxycholic acid (CDCA) ratio in maternal and cord blood plasma. This indicates a continued dysregulation of bile acid metabolism despite therapeutic intervention. Maternal plasma lipid analysis showed a distinct maternal dyslipidemia pattern among ICP patients, marked by elevated cholesterol levels on VLDL particles and heightened triglyceride concentrations on LDL particles, persisting even after UDCA treatment. Cord plasma lipid profiles in ICP patients exhibited elevated triglyceride and free fatty acid levels alongside a tendency toward increased β-hydroxybutyrate. The changes in lipid metabolism in both maternal and cord blood correlated with the high CA/CDCA ratio, but not total bile acid levels or gestational diabetes status. Understanding the imbalances in maternal and cord bile acid and lipid profiles that persist after standard UDCA therapy provides insights for improving management strategies and mitigating the long-term consequences of ICP.

**News and Noteworthy:** This study uncovers that despite ursodeoxycholic acid treatment, intrahepatic cholestasis of pregnancy (ICP) is associated with increases in the ratio of cholic acid to chenodeoxycholic acid in both maternal and cord blood, suggesting ongoing dysregulation of bile acid metabolism. The high cholic to chenodeoxycholic acid ratio is correlated with maternal dyslipidemia and high cord blood lipids. These findings may inform more targeted approaches to managing ICP.

## Introduction

Intrahepatic cholestasis of pregnancy (ICP) is a condition characterized by elevated plasma bile acids (BAs) and liver transaminases, maternal pruritus, and adverse pregnancy outcomes (1). Susceptibility to ICP is influenced by genetic variation (2–13) and steroid hormones, which inhibit BA transport through multiple mechanisms (14). The latter occurs to a mild extent in normal pregnancy (15–17), and there is evidence that maternal circulating BAs are subclinically elevated in normal pregnancy (14). In ICP, these effects may be exacerbated due to genetic variation in BA transporters (2–13), and due to increases in certain hormone products such as sulfated progesterone metabolites, which are elevated in ICP patients (18–20). These metabolites appear to be cholestatic due to a combination of multiple mechanisms: (i) they competitively inhibit BA binding and transport by Na^+^-taurocholate transporting polypeptide (NTCP), the sinusoidal uptake transporter for BAs (21); (ii) they can trans-inhibit the bile salt export pump (BSEP), the canalicular efflux transporter for BAs (22); and (iii) they also competitively inhibit BA binding and activation of the farnesoid X receptor (FXR), the BA-regulated nuclear receptor that transcriptionally induces hepatobiliary BA efflux (23).

In addition to high maternal serum BAs, there are other notable alterations in BAs during ICP. First, cord serum BAs are also elevated (24, 25). Second, there are characteristic alterations in BA composition. The most well-recognized change, which occurs in both maternal and cord serum, is an increase in the ratio of cholic acid (CA) to chenodeoxycholic acid (CDCA), the two primary BAs synthesized by the human liver (18, 24, 26–28).

The most common treatment for ICP is ursodeoxycholic acid (UDCA), which improves liver function tests and reduces pruritus (29). In some studies, UDCA also reduces maternal total BAs (24, 29), although this has not been observed in all trials (30, 31). UDCA has also been reported to partially reverse the qualitative changes in BA composition and maternal and cord serum (18, 24), though other reports indicate that UDCA does not restore cord BA composition (32).

ICP is also associated with alterations in maternal and fetal metabolic health: it has been reported that mothers with the condition are more likely to be diagnosed with gestational diabetes (33). They also show altered lipid profiles. Some reports show higher triglycerides, higher low-density lipoprotein cholesterol (LDL-C), and lower high-density lipoprotein cholesterol (HDL-C) (34–38), although this has not been fully characterized using detailed lipoprotein fractionation methods. Other reports indicate that ICP, like other cholestatic conditions, results in the accumulation of Lipoprotein X (39, 40). Lipoprotein X has a similar density to LDL, but has a similar size to VLDL, and it is particularly rich in phospholipids and free cholesterol. Altered maternal lipids are known to be associated with adverse pregnancy outcomes (41). Children born to mothers with ICP have been reported to be more likely to have fetal growth abnormalities, including being born large for gestational age (42) or having fetal growth restriction (43, 44) and to show metabolic abnormalities during adolescence (45). In a mouse model of maternal hypercholanemia–wherein pregnant dams were fed a diet containing CA–female offspring were more susceptible to western diet-induced weight gain, glucose intolerance, and higher cholesterol in plasma and liver (45). This suggests metabolic reprogramming as a consequence of the *in utero* environment. However, fetal metabolic features during ICP, or ICP with gestational diabetes, are incompletely characterized.

BAs are known to regulate glucose and lipid metabolic parameters, through their physicochemical role in lipid absorption and through their ability to activate BA receptors such as FXR and TGR5 (46–48). BA signaling through FXR in the maternal liver and FXR in the placenta or fetal liver could potentially affect lipid metabolism in both mother and fetus. Moreover, data from preclinical models, and emerging genetic evidence in humans, suggest that BA composition–especially as it relates to cholic acid–also influences metabolic phenotypes (49, 50). However, the role of BA composition in the development of the metabolic effects of ICP are unknown. The aims of this study were to (i) characterize the maternal and cord blood metabolic features in UDCA-treated patients with ICP alone or both ICP and gestational diabetes; and (ii) determine the associations of these metabolic parameters with BA levels and BA composition.

## Materials and methods

### Subjects

We recruited 20 women pregnant with singletons: 7 ICP patients without diabetes (ICP-NoDiab) and 6 ICP patients with gestational diabetes (ICP-Diab), who had a scheduled delivery between 36 and 37 weeks (according to the clinical recommendations at the time of the study); and 7 control subjects without ICP, who were also scheduled to deliver in the same time frame. Women in the ICP-Diab group were diagnosed with gestational diabetes before the onset of cholestasis, and their diabetes care was managed by their physicians. The racial distribution of each group was: controls (3 white, 4 other), ICP-NoDiab (1 Asian, 3 white, 2 other), and ICP-Diab (1 Asian, 5 other). Ethnicity distribution was: controls (3 Hispanic, 3 Non-Hispanic, 1 other), ICP-NoDiab (4 Hispanic, 2 Non-Hispanic), ICP-Diab (5 Hispanic, 1 other). Exclusion criteria for recruitment included chronic hepatobiliary disease, multiple gestations, maternal systemic lupus erythematosus, and fetuses with identified heart block or suspected major congenital anomalies. One additional subject who had ICP without diabetes (patient #21) underwent our testing protocol, but was excluded from the group analyses, because she had previously undergone Roux-en-Y gastric bypass surgery, which is known to influence glucose and lipid metabolism (51), as well as BA levels and composition (52–56). We report the data from this subject in the online supplementary data (Supplementary Table S3). ICP was defined by pruritus and BAs >10 μmol/l. All ICP patients were prescribed UDCA by their physicians at the time of diagnosis. Fasting maternal blood was collected 3-5 days prior to delivery. Cord blood was collected at delivery. Body Mass Index (BMI) was available for women whose physicians had previously collected the information, early in the pregnancy. This study was approved by the institutional review board of Columbia University Medical Center and informed consent was received from all patients.

### BA measurements

Fasting BA and alanine aminotransferase (ALT) measurements at the time of diagnosis were carried out by ARUP laboratories. For BA measurements in fasting maternal and cord plasma collected for full analysis, measurements were carried out at the Biomarkers Core Laboratory at Columbia’s Irving Institute for Clinical and Translational Research. Briefly, 100 μl plasma was combined with 10 μl internal standard (cholate-d4 50nM, ISOTECH) and 1 ml ice-cold acetonitrile. Samples were vortexed, centrifuged for 15 minutes at 11,000xg, and supernatants were transferred to ultraperformance liquid chromatography-tandem mass spectrometry (UPLC-MS/MS) vials. Samples were dried at 45° C under nitrogen, resuspended in 100 μl 55%/45% (v/v) methanol/water (both with 5 mM ammonium formate). 5 μl were injected into UPLC-MS/MS. Samples were compared against working standards of each BA species measured. BA levels are reported in μmol/l.

### Hormone and metabolite measurements

The fasting blood samples were also used to perform hormone and metabolite measurements. Plasma insulin and C-peptide were measured using ELISA kits (Mercodia), and glucose was analyzed with a hexokinase kit (Glucose HK, Sigma). Lipids were measured using colorimetric assays: triglycerides (Infinity, Thermo Scientific), free fatty acids, cholesterol, free cholesterol and phospholipids (Wako Diagnostics). Cholesteryl ester was calculated by subtracting free cholesterol from total cholesterol. β-Hydroxybutyrate and ALT were measured with assay kits (β-Hydroxybutyrate, Liquicolor Stanbio and ALT reagent, Teco Diagnostic). FGF19 was measured by ELISA (R&D Systems).

### Lipoprotein fractionation

VLDL (d<1.006 g/mL), LDL (1.006<d<1.063 g/mL) and HDL fractions (1.063<d<1.210 g/mL) were separated by sequential density ultracentrifugation, using the Optima MAX-TL Ultracentrifuge with TLA-100 rotor (Beckman Coulter). Fast protein liquid chromatography (FPLC) was carried out by running 200 μl of plasma onto a Superose6 10/300 GL column (Amersham Pharmacia Biotech), and fractions were collected using the fraction collector FC-204 (Gilson). Fractions were analyzed with colorimetric kits as mentioned above.

### Statistical analyses

All analyses were performed in the open source statistical package R. Data are reported as mean ± standard error, or median [interquartile range], for normally or non-normally distributed data, respectively. For comparisons of non-normally distributed data between two groups, we used Mann-Whitney tests, or between three groups, we used Kruskall-Wallis and Dunn posthoc tests. For paired comparison before and after UDCA treatment, we used Wilcoxon signed rank test. Spearman rank correlations were calculated with non-transformed data.

## Results

### Subject characteristics

Data are shown in **Table 1**. There were no differences between groups in maternal age, gestational age at maternal blood collection, gestational age at delivery, or birthweight. Among the ICP patients, there was no effect of diabetes on gestational age at initiation of UDCA treatment, highest total BAs (as measured by ARUP clinical laboratories), or highest ALT. Seven ICP patients (3 ICP-NoDiab and 4 ICP-Diab) had highest total BAs above 40 μmol/l, which is considered severe ICP (57).

**Table 1.** Patient characteristics.

|  | **Control** | **ICP-all** | **ICP-NoDiab** | **ICP-Diab** |
| --- | --- | --- | --- | --- |
| **N** | 7 | 12 | 6 | 6 |
| **Maternal age (years)** | 34.1 ± 1.3 | 32.4 ± 2.3 | 30.2 ± 3.5 | 34.7 ± 2.8 |
| **BMI (kg/m<sup>2</sup>) †** | 26.6 ± 2.1 | 28.4 ± 2.0 |  |  |
| **Highest total BAs–ARUP labs (μmol/l)** | - | 56 [36.7] | 42 [39.3] | 59.9 [33.5] |
| **# patients with BAs–ARUP labs >40 μmol/l** | - | 7 (58%) | 3 (50%) | 4 (67%) |
| **Highest ALT–ARUP labs (IU/l)** | - | 123 [267.5] | 120.5 [200] | 124 [326] |
| **GA at initiation of UDCA therapy (wks)** | - | 34.2 ± 0.7 | 33.9 ± 0.8 | 34.5 ± 1.1 |
| **GA at maternal blood collection (wks)** | 36.6 ± 0.2 | 36.6 ± 0.1 | 36.7 ± 0.2 | 36.4 ± 0.2 |
| **GA at delivery (wks)** | 37.0 ± 0.1 | 37.3 ± 0.2 | 37.2 ± 0.1 | 37.5 ± 0.3 |
| **Birthweight (g)** | 2827 ± 94 | 3068 ± 147 | 3095 ± 278 | 3040 ± 132 |

†BMI was available for 6/7 controls and 7/12 ICP patients and was determined at GA 10.0 ± 1.5 weeks. Data is presented as the median ± [interquartile range].
Abbreviations: ALT=alanine aminotransferase; BMI=body mass index; GA=gestational age

### Maternal bile acid analysis

Total maternal fasting BAs were high at diagnosis and markedly decreased in most ICP patients after UDCA treatment (**Fig. 1A**), such that there were no significant differences in total BAs between UDCA-treated ICP patients and controls (**Fig. 1C**). These findings are consistent with the effect of UDCA to lower plasma BAs. Complete BA data is listed in **Supplementary Table S1.**

**Figure 1.**
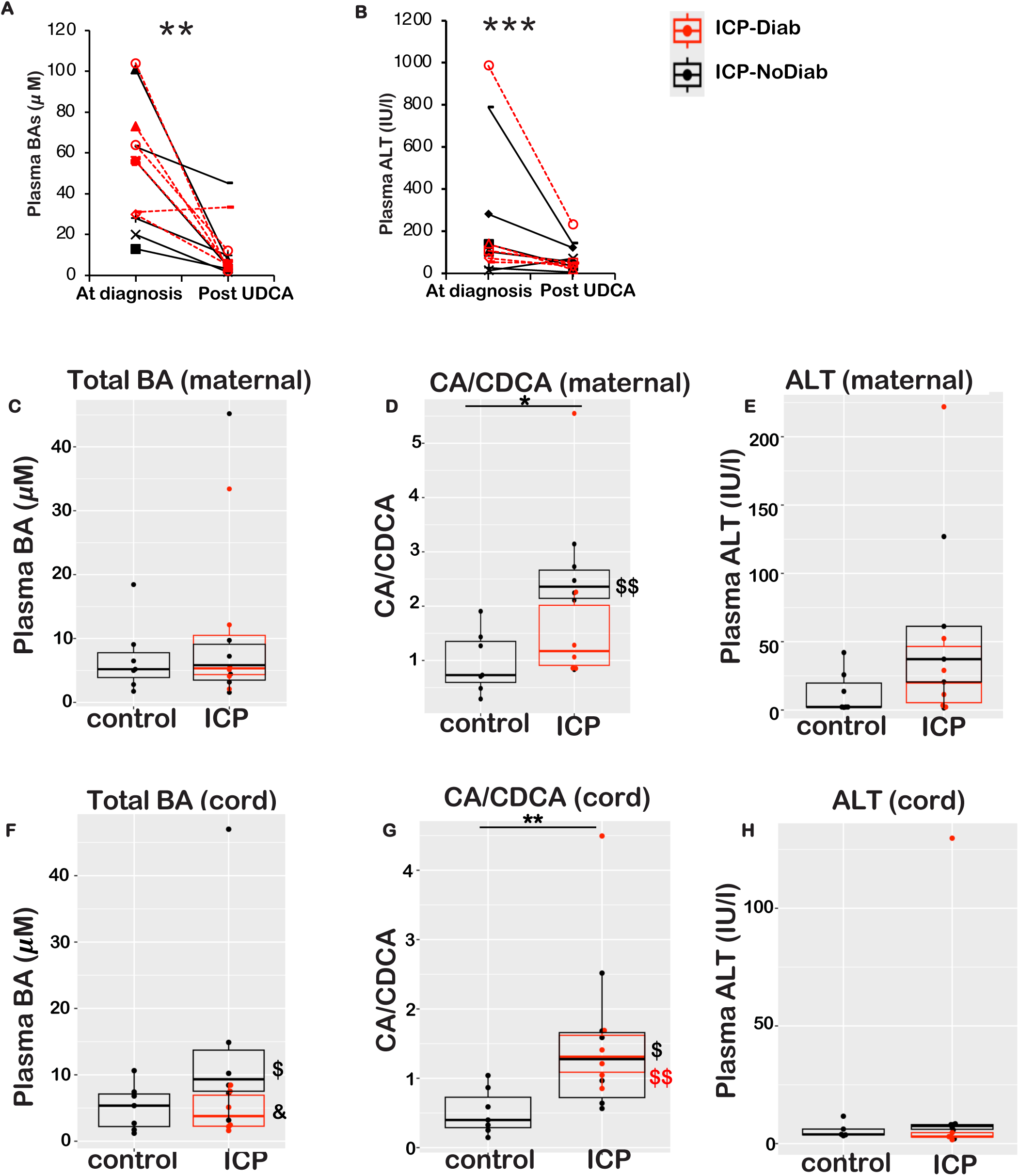
Bile acid levels and composition. (A-B) Total plasma BAs and ALT in ICP-diagnosed individuals at the time of diagnosis and initiation of UDCA treatment, and ∼2.7 weeks later, at the time of fasting maternal blood collection for full analysis. The red lines represent the ICP-Diab group and the black lines represent the ICP-NoDiab group. (C) Total fasting plasma BAs in all maternal subjects at the time of maternal blood collection. (D) Ratio of cholic acid (CA) and its conjugated forms to chenodeoxycholic acid (CDCA) and its conjugated forms. (E) Plasma ALT in all maternal subjects at the time of maternal blood collection. (F) Total BAs in cord blood plasma. (G) Ratio of CA and its conjugated forms to CDCA and its conjugated forms, in cord blood plasma. (H) ALT in cord blood plasma. Data show mean ± SEM. *P<0.05, **P<0.01, ***P<0.001 for ICP-all (n=12) versus controls (n=7) by Wilcoxon signed rank test (for panels A-B) or Mann-Whitney test (for panels C-H). ^$^P<0.05, ^$$^P<0.01 for ICP-NoDiab (n=6) or ICP-Diab (n=6) versus controls (n=7) and ^&^P<0.05 for ICP-Diab (n=6) versus ICP-NoDiab (n=7) by Kruskall-Wallis test with posthoc Dunn tests.

Next, we examined differences in BA composition between groups. Consistent with prior reports (18, 24, 26–28), we found that ICP patients had an increase in the ratio of CA to CDCA (including conjugated and unconjugated forms of each) (**Fig. 1D**). A depiction of the relative concentrations of individual BAs between groups is shown in **Supplementary Figure S1A**, and absolute values, median, and interquartile range are provided in **Supplementary Table S1**. These data suggest that in ICP patients, even after total BAs are normalized by UDCA treatment, the CA/CDCA ratio remains elevated.

As expected ALT was decreased by UDCA treatment (30), (**Fig. 1B**), such that at the time of maternal blood collection for full analyses, there were no differences between the groups (**Fig. 1E**). Next, we measured FGF19 as a marker of FXR activity. FXR regulates BA synthesis through the inhibitory action of FGF19 on the expression of the BA synthesis enzyme CYP7A1. FGF19 was highly variable and not significantly different between groups (median [interquartile range] for control and ICP, respectively, were 91.1 [81.9] and 53.4 [53.4] pg/ml, P = 0.22).

### Cord bile acid analysis

We observed no significant difference in total BAs from cord blood plasma between the controls and combined ICP groups (**Fig. 1F** and **Supplementary Table S2**). However, ICP-NoDiab had higher cord BAs than those in the control or ICP-Diab groups. The BA composition of cord plasma was markedly different than maternal plasma, in that the pool was almost entirely composed of conjugated, primary BAs, with unconjugated and secondary species each making up less than 2% of the total pool (**Supplementary Table S2**).

The CA/CDCA ratio was increased threefold in cord plasma from ICP groups compared to controls (**Fig. 1G** and **Supplementary Fig. S1B**). There were no differences in ALT between groups (**Fig. 1H**). FGF19 was highly variable and not significantly different between groups (median [interquartile range] for control and ICP, respectively, were 19.2 [15.8] and 51.9 [28.5] pg/ml, P = 0.14).

### Glycemic parameters

As a whole group, there were no differences in fasting plasma glucose levels between controls and ICP-Diab or combined ICP groups (**Table 2**). However, there were lower levels of glucose, insulin, C-peptide, and HOMA-IR in ICP-NoDiab compared to ICP-Diab or controls.

**Table 2.**
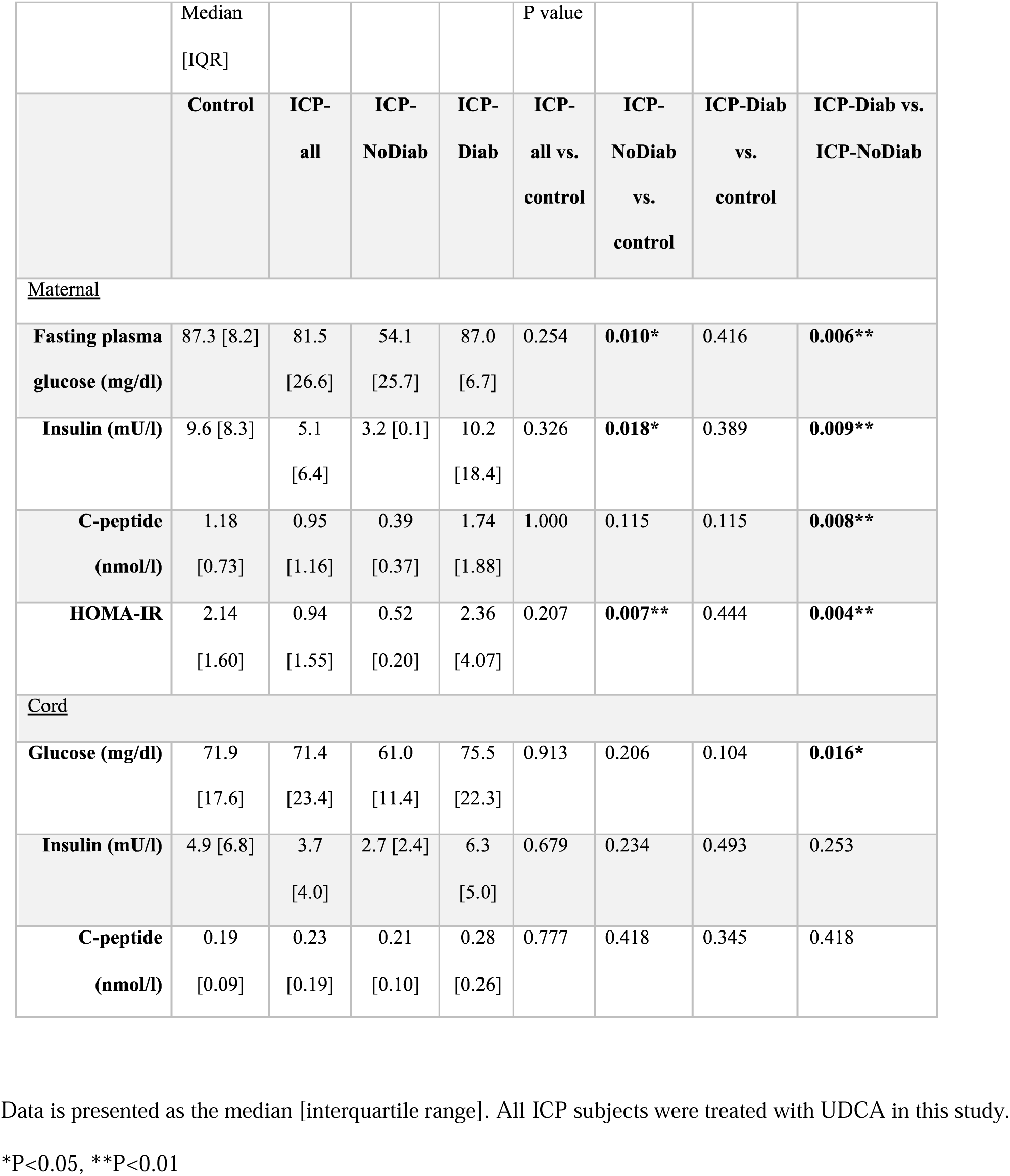
Glycemic parameters.

Cord plasma glycemic parameters were all substantially lower than maternal parameters (**Table 2**). Moreover, they were similar across groups. The only difference we observed was an increase in the plasma glucose in the ICP-Diab group compared to the ICP-NoDiab group.

### Maternal lipid analysis

In maternal plasma, there were no significant differences between groups in total cholesterol, total triglycerides, or free fatty acids (**Fig. 2A-C**). However, ICP-NoDiab subjects had an increase in levels of the ketone body β-hydroxybutyrate (**Fig. 2D**).

**Figure 2.**
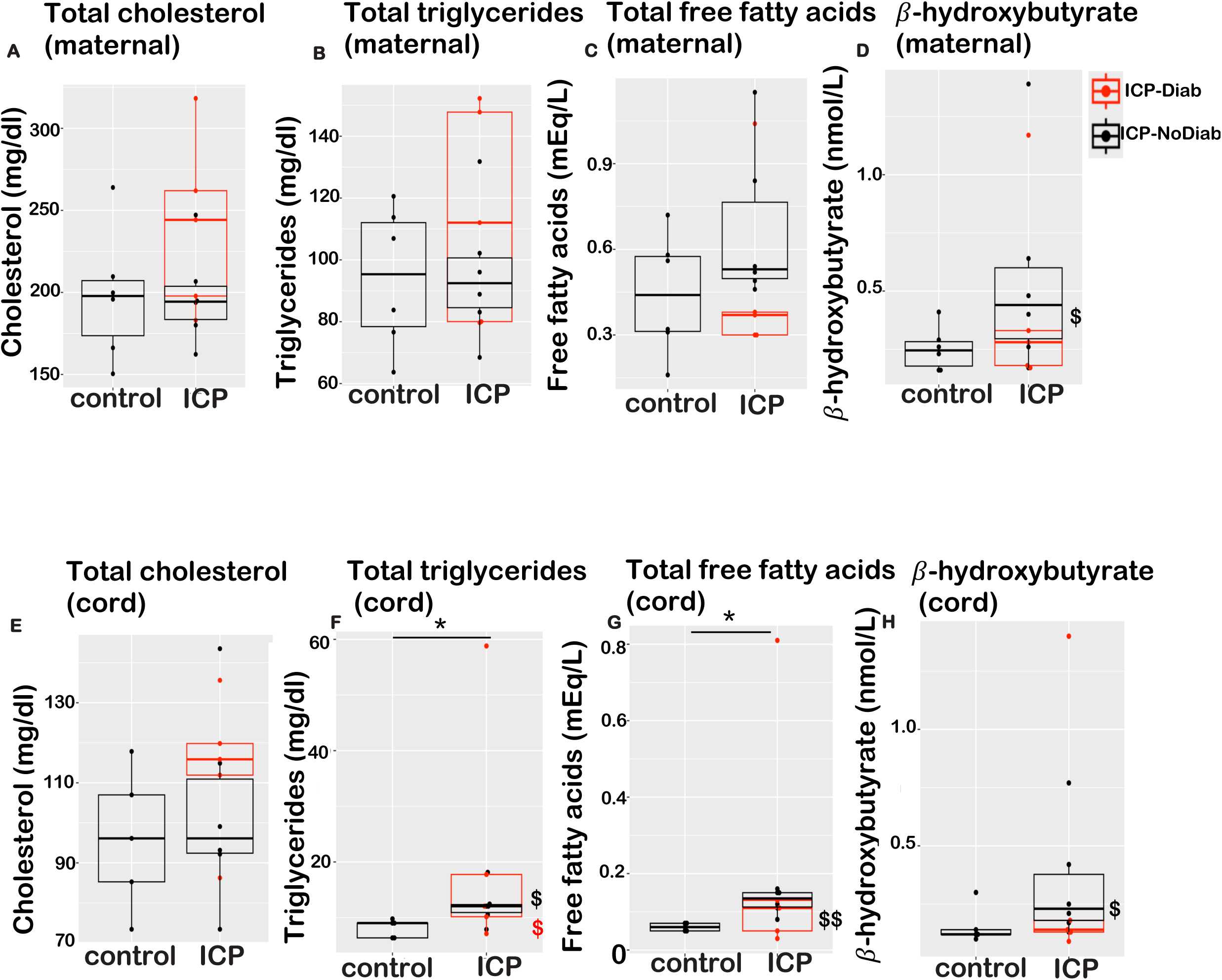
Total plasma lipids. (A) Total cholesterol, (B) total triglycerides, (C) free fatty acids, and (D) β-hydroxybutyrate in maternal plasma. (E) Total cholesterol, (F) total triglycerides, (G) free fatty acids, and (H) β-hydroxybutyrate in cord blood plasma. The box plot shows the distribution of the dataset with a focus on the median value *P<0.05 for ICP-all (n=12 versus controls (n=7) by Mann-Whitney test. ^$^P<0.05, ^$$^P<0.01 for ICP-NoDiab (n=6) or ICP-Diab (n=6) versus controls (n=7) by Kruskall-Wallis test with posthoc Dunn tests.

Next, we examined lipoprotein distribution by sequential density ultracentrifugation and fast protein liquid chromatography. We found that ICP patients had increased VLDL-cholesterol, which was mostly cholesteryl ester (**Fig. 3A-C, Supplementary Fig. 2A**). ICP subjects also had high LDL-triglycerides (**Fig. 3D, Supplementary Fig. 2B**). ICP-Diab patients also showed elevated VLDL-triglycerides and phospholipids compared to controls (**Fig. 3D-E**). For comparison to prior studies, we also analyzed maternal HDL-cholesterol using a commercial kit relying on precipitation of apoB-containing lipoproteins, then calculated the LDL-cholesterol using the Friedewald formula (58). In this analysis, we observed no significant differences between groups in HDL-cholesterol (mean values 51.8 ± 8.8, 34.4 ± 6.1, 33.7 ± 5.2 mg/dl for control, ICP-NoDiab, and ICP-Diab, respectively; all P>0.05) or LDL-cholesterol (mean values 127.0 ± 17.6, 144.1 ± 9.7, 184.5 ± 27.5 mg/dl for control, ICP-NoDiab, and ICP-Diab, respectively; all P>0.05).

**Figure 3.**
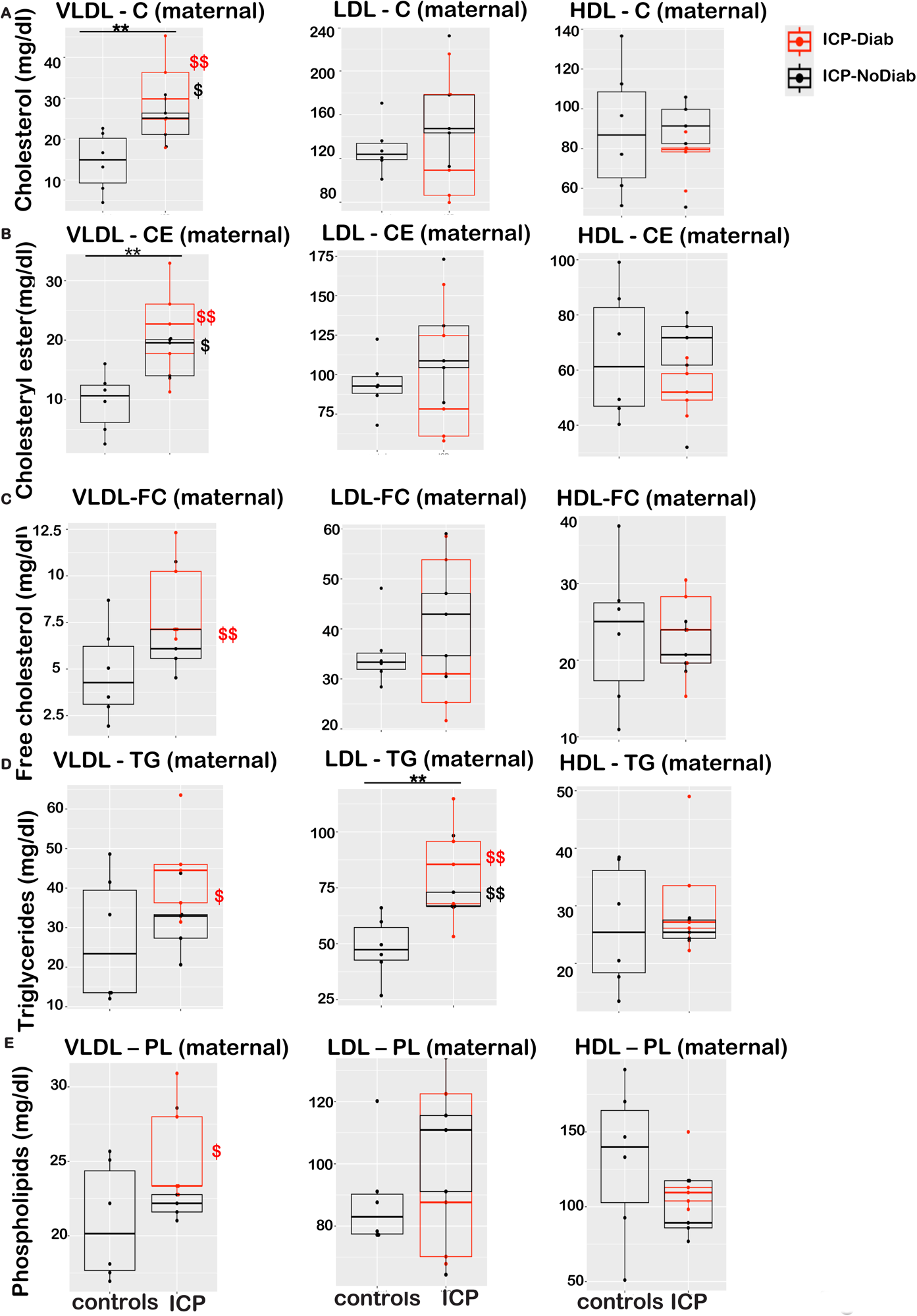
Lipids in fractionated maternal plasma. (A-E) Very-low-density lipoprotein (VLDL) (d<1.006), low-density lipoprotein (LDL) (1.006<d<1.063), and high-density lipoprotein (HDL) (1.063<d<1.210) fractionated by sequential density ultracentrifugation. (A) C=cholesterol, (B) CE=cholesteryl ester, (C) FC=free cholesterol, (D) TG=triglycerides, (E) PL=phospholipid. The box plot shows the distribution of the dataset and the line within the box represents the median value**P<0.01 for ICP-all (n=12) versus controls (n=7) by Mann-Whitney test. ^$^P<0.05, ^$$^P<0.01 for ICP-NoDiab (n=6) or ICP-Diab (n=6) versus controls (n=7) by Kruskall-Wallis test with posthoc Dunn tests.

### Cord lipid analysis

Cord plasma lipids were generally lower than those from maternal plasma (**Fig. 2E-H**), as expected (59, 60). Total plasma cholesterol–which arises from delivery of maternal cholesterol across the placenta and *de novo* synthesis in the fetus (61–63)–was not different between groups (**Fig. 2E**). Total triglycerides and free fatty acids arise from placental transfer of maternal free fatty acids and fatty acids derived from maternal triglyceride-rich remnant particles, as well as *de novo* synthesis (59, 64). We found that cord plasma triglycerides and free fatty acids were two- to three-fold higher in the ICP groups (**Fig. 2F-G**). Ketone bodies are a fuel source and lipogenic precursor in the fetus, but cannot be synthesized by the fetus itself, thus are supplied exclusively by diffusion across the placenta from maternal plasma (60, 61). Consistent with this, we found β-hydroxybutyrate levels were similar in cord and maternal plasma (**Fig. 2H**, compare to Fig. 2D). Compared to controls, β-hydroxybutyrate was higher in the ICP-NoDiab group, and the ICP-Diab group showed a similar trend (**Fig. 2H**). There were no significant differences in these cord lipid parameters between the ICP-NoDiab and ICP-Diab groups.

The distribution of cholesterol across lipoprotein fractions in cord plasma was different than maternal plasma, with substantially more of the cholesterol residing on HDL particles, consistent with prior reports (**Fig. 4A and Supplementary Fig. 2C**) (59). The levels of cholesterol on LDL particles in cord plasma is likely determined by a combination of several pathways: (i) secretion of apoB-containing particles from fetal liver (ii) and placenta (65), (iii) uptake in liver, as LDL-cholesterol levels are tightly inversely correlated with fetal liver LDL receptor (66), and (iv) uptake in adrenals, which convert cholesterol into steroid hormones at a rate 5- to 10-fold higher than in adults (67). HDL particles can be secreted from the fetal-facing membrane of placental trophoblasts, and cholesterol can also be effluxed onto HDL or apoA-I from trophoblasts and from endothelial cells of the fetoplacental vasculature (62, 63, 68). The ICP-Diab group showed higher free cholesterol on VLDL and lower cholesterol and free cholesterol on LDL and HDL, compared to controls, but other cholesterol distribution parameters were not different between groups (**Fig. 4A-C**). Lipoprotein association of triglycerides was also different in cord compared to maternal plasma, with more triglycerides residing on LDL particles (**Fig. 4D and Supplementary Fig. 2D**) (69). We observed no significant differences in triglyceride or phospholipid distribution between groups.

**Figure 4.**
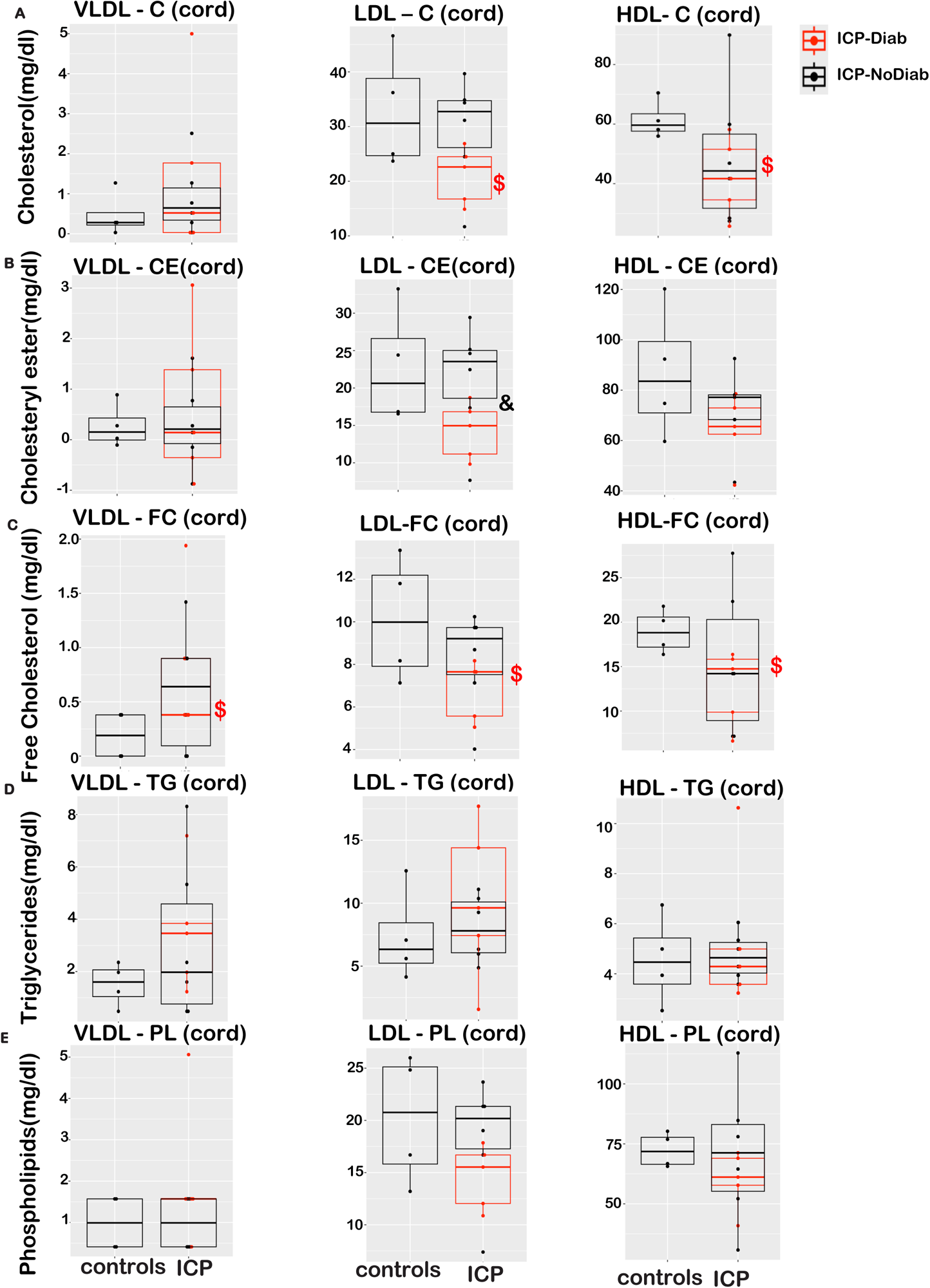
Lipids in fractionated cord blood plasma. (A-E) VLDL (d<1.006), LDL (1.006<d<1.063), and HDL (1.063<d<1.210) fractionated by sequential density ultracentrifugation. (A) C=cholesterol, (B) CE=cholesteryl ester, (C) FC=free cholesterol, (D) TG=triglycerides, (E) PL=phospholipid. The box plot shows the distribution of the dataset and the line within the box represents the median value.^$^P<0.05 for ICP-NoDiab (n=6) or ICP-Diab (n=6) versus controls (n=7) and ^&^P<0.05 for ICP-Diab versus ICP-NoDiab by Kruskall-Wallis test with posthoc Dunn tests.

### Correlations between BAs and metabolic parameters

We investigated the relationships between maternal BAs and the metabolic features of the mothers that were altered in ICP, using Spearman rank correlations. We found that the maternal CA/CDCA ratio–but not maternal total BAs–was significantly positively correlated with maternal ALT, VLDL-cholesterol, and β-hydroxybutyrate (**Table 3**).

**Table 3.** Spearman rank correlations. Correlations between the metabolic parameters that were altered in ICP, and total BAs or the CA/CDCA ratio.

|  |  | All |  |
| --- | --- | --- | --- |
| BA parameter | Metabolic parameter | R<sub>s</sub> | P |
| Maternal BAs–maternal metabolites |  |  |  |
| Total BAs | ALT | 0.35 | 0.163 |
| Total BAs | VLDL-cholesterol | 0.26 | 0.322 |
| Total BAs | LDL-triglyceride | -0.11 | 0.676 |
| Total BAs | βOH butyrate | 0.26 | 0.308 |
| CA/CDCA | ALT | 0.78 | **0.0002\*\*\*** |
| CA/CDCA | VLDL-cholesterol | 0.51 | **0.046\*** |
| CA/CDCA | LDL-triglyceride | 0.26 | 0.332 |
| CA/CDCA | βOH butyrate | 0.60 | **0.011\*** |
| Cord BAs–cord metabolites |  |  |  |
| Total BAs | Triglyceride | 0.42 | 0.108 |
| Total BAs | Free fatty acids | 0.35 | 0.183 |
| Total BAs | βOH butyrate | 0.21 | 0.437 |
| CA/CDCA | Triglyceride | 0.62 | **0.010\*** |
| CA/CDCA | Free fatty acids | 0.44 | 0.085 |
| CA/CDCA | βOH butyrate | 0.07 | 0.807 |
| Maternal BAs–cord metabolites |  |  |  |
| Total BAs | Triglyceride | 0.48 | 0.058 |
| Total BAs | Free fatty acids | 0.09 | 0.740 |
| Total BAs | βOH butyrate | 0.03 | 0.901 |
| CA/CDCA | Triglyceride | 0.61 | **0.012\*** |
| CA/CDCA | Free fatty acids | 0.56 | **0.023\*** |
| CA/CDCA | βOH butyrate | 0.35 | 0.185 |

\*P<0.05, \*\*\*P<0.001

We next investigated the associations of BAs on the cord plasma parameters that were elevated in ICP. We found that the cord CA/CDCA ratio–but not cord total BAs–was positively correlated with cord triglycerides (**Table 3**). Interestingly, we also found that the maternal CA/CDCA ratio was positively correlated with cord triglycerides and free fatty acids (**Table 3**).

## Discussion

The major findings of this work are that ICP is associated–even after UDCA therapy–with (i) higher CA/CDCA ratio in maternal and cord plasma; (ii) a unique maternal dyslipidemia with high cholesterol on VLDL particles and high triglycerides on LDL or remnant particles; (iii) two- to three-fold higher cord triglycerides and free fatty acids; and (iv) a tendency towards higher ketones. Finally, we found that (v) additional diagnosis of gestational diabetes does not strongly further impact the BA and lipid phenotypes of ICP.

An increase in the CA/CDCA ratio has been noted in ICP patients (18, 24, 26–28). Our data suggests that this alteration in BA composition persists after UDCA therapy. The cause of this change is unknown. One potential hypothesis arises from the known accumulation of sulfated progesterone metabolites, which competitively inhibit BAs’ ability to activate their receptor FXR (23). This mechanism has been invoked to explain reduced expression of BA transporters (23), which are normally induced by FXR. Another effect of FXR is to suppress the expression of *CYP8B1*, the hepatic enzyme responsible for determining the ratio of CA/CDCA (70). Thus, it is possible that FXR inhibition by sulfated steroid hormones, such as sulfated progesterone metabolites (23) can release this suppression, consequently inducing preferential CA production. FXR also normally suppresses *CYP7A1*, the rate limiting enzyme of BA synthesis (71, 72), but CYP7A1 activity, as measured by the validated plasma marker of its activity (“C4” or 7α-hydroxy-4-cholesten-3-one), is actually decreased in ICP patients before and after UDCA treatment (20). Perhaps CYP8B1 is uniquely regulated by a certain subset of sulfated progesterones, or other steroid hormones altered in ICP (73).

The metabolic sequelae of ICP have received relatively less attention. Prior studies have used chemical precipitation of HDL or LDL particles to report that ICP patients have a tendency towards high total triglycerides, high LDL-cholesterol and low HDL-cholesterol (34–38). On the other hand, other evidence has suggested that cholestasis, including ICP, is associated with accumulation of Lipoprotein X (39, 40). Here, we used ultracentrifugation and size-exclusion chromatography to directly separate lipoproteins by density and particle size. These analyses revealed that ICP is associated with high VLDL-cholesterol, and high triglycerides on particles the size and density of LDL or remnants. This is a unique dyslipidemia, and suggests the possibility that in ICP–potentially through altered steroid hormone or BA signaling–affects either (i) the composition of secreted apoB-containing lipoprotein particles or (ii) the partitioning of lipoprotein clearance from plasma. It is also possible that ICP and lipoprotein traits have some shared genetic underpinnings, as common variants in several loci achieve genome-wide significance for both (74, 75).

The source of the increased cord plasma triglycerides and free fatty acids is unknown. Because these lipids arise partly from transfer of fatty acids derived from maternal triglyceride-rich lipoproteins across the placenta (59, 64), and the ICP patients showed high LDL-triglycerides, this may suggest a link. Another source of these lipids is from cord *de novo* synthesis (59, 64), partly from ketone body precursors (60, 61). Indeed, the ketone body β-hydroxybutyrate tended to be elevated in ICP patients and cord plasma. This suggests the possibility that alterations in maternal lipid metabolism during ICP may cause the high triglycerides and free fatty acids in cord blood plasma.

BAs regulate multiple aspects of lipid metabolism, and many of these effects occur through activation of the nuclear receptor FXR. This includes the effect of FXR activation to lower plasma triglycerides (76). Studies in mice also demonstrate effects of FXR activation to lower free fatty acids and β-hydroxybutyrate (77). It is of interest whether impaired FXR activity contributes to the lipid phenotypes during ICP. One possibility is impaired activation of FXR due accumulation of sulfated progesterone metabolites, which are elevated in ICP and which are known to competitively inhibit FXR (23). Another possibility is that the change in BA composition, which exists even after treatment, causes differential FXR activity. A robust change in BA composition in ICP, which occurs in maternal and cord blood, is the increase in CA and the CA/CDCA ratio. Because CA is a weaker activator of human FXR compared to CDCA (78–80), it raises the possibility that the elevated CA/CDCA ratio reduces FXR activity in ICP patients. Supporting a possible role for this effect in the lipid phenotypes of ICP, we observed significant correlations between maternal and cord CA/CDCA ratios with plasma lipid levels.

ICP and gestational diabetes may be linked, as women diagnosed with ICP are reported to be at an increased risk of gestational diabetes (33). Prior studies have shown that gestational diabetes is associated with alterations in maternal lipoprotein metabolism, including increased triglycerides and increased levels of small dense LDL (41). Our finding that gestational diabetes did not contribute additional defects in maternal or cord lipids beyond the effects of ICP suggests that either ICP has a more dominant effect on lipid metabolism, or that the defects in lipid metabolism in ICP and gestational diabetes arise from shared mechanisms.

The strengths of this study include: (i) assessment of metabolic parameters in both maternal and cord blood; (ii) the use of additional methods of lipoprotein fractionation by size (liquid chromatography) and density (ultracentrifugation), in comparison to previous publications that used precipitation methods (34, 37, 38); and (iii) inclusion of ICP patients with and without gestational diabetes. Limitations include: (i) small sample size; (ii) cross-sectional design; and (iii) lack of data from ICP patients prior to UDCA treatment.

In this work we established that ICP is linked to alterations in BA composition in maternal and cord blood, which persist after UDCA treatment, despite resolution of the high total BAs and ALT. We hypothesize that the altered CA/CDCA ratio could contribute to lipid alterations, including high levels of triglycerides and free fatty acids in cord blood, and high levels of β-hydroxybutyrate and altered lipoprotein composition in maternal blood. These alterations were correlated to the CA/CDCA ratio, but not total BAs, in both maternal and cord blood plasma. This suggests the possibility that altered BA composition is an underappreciated contributor to the metabolic alterations associated with ICP.

## SUPPLEMENTAL MATERIAL

Supplemental Figs. S1 and S2: https://doi.org/10.6084/m9.figshare.26738695.v2

Supplemental Tables S1–S3: https://doi.org/10.6084/m9.figshare.26738695.v2

## Data availability

All source data will be made available upon request to the corresponding author.

## Data Availability

All data produced in the present study are available upon reasonable request to the authors.

## Acknowledgements/grant support

This work was supported by funding from the National Institutes of Health R01DK115825, R01DK135298, and R01HL125649 and the Russell Berrie Foundation, and funding from an American Medical Association seed grant (to SGO). This publication was also supported by the National Center for Advancing Translational Sciences, National Institutes of Health, through Grant Number UL1TR001873 and the National Institute of Diabetes and Digestive and Kidney Diseases, through grants P30DK132710 and P30DK063608. The content is solely the responsibility of the authors and does not necessarily represent the official views of the NIH.

## Abbreviations

ALT: alanine aminotransferase
BAs: bile acids
BMI: body mass index
BSEP: bile salt export pump, the canalicular BA efflux transporter
CA: cholic acid
CDCA: chenodeoxycholic acid
CE: cholesteryl ester
FC: free cholesterol
FPLC: fast protein liquid chromatography
GA: gestational age
HDL-C: high-density lipoprotein-cholesterol
ICP: intrahepatic cholestasis of pregnancy
ICP-diab: ICP plus diabetes
ICP-NoDiab: ICP without diabetes
LDL-C: low-density lipoprotein-cholesterol
NTCP: sodium-taurocholate cotransporting peptide, a sinusoidal BA uptake transporter
PL: phospholipid
TG: triglyceride
UDCA: ursodeoxycholic acid

