## Supplementary figures and images for "Unresolved alterations in bile acid composition and dyslipidemia in maternal and cord blood after ursodeoxycholic acid treatment for intrahepatic cholestasis of pregnancy"

### Supplemental Figure 1 and 2

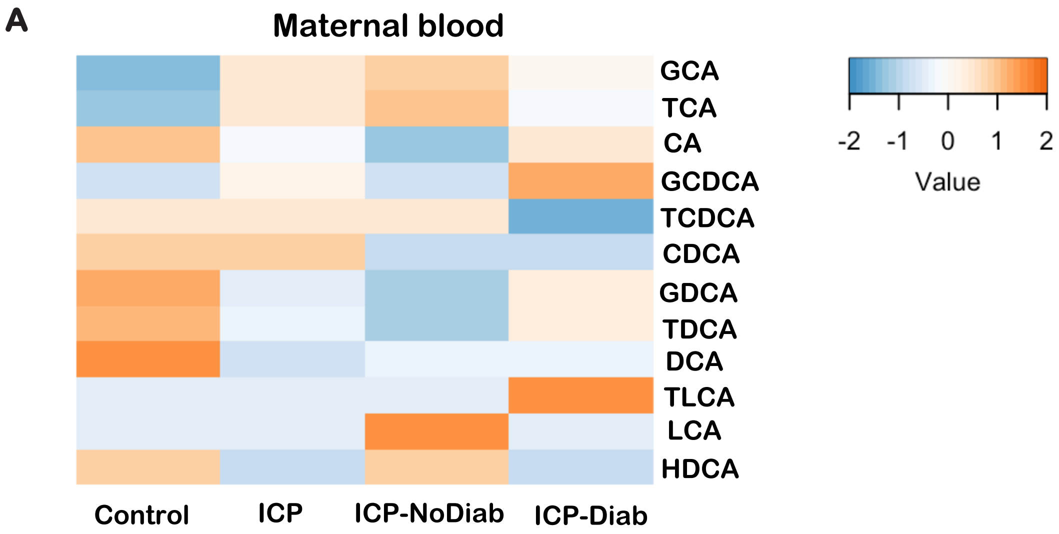


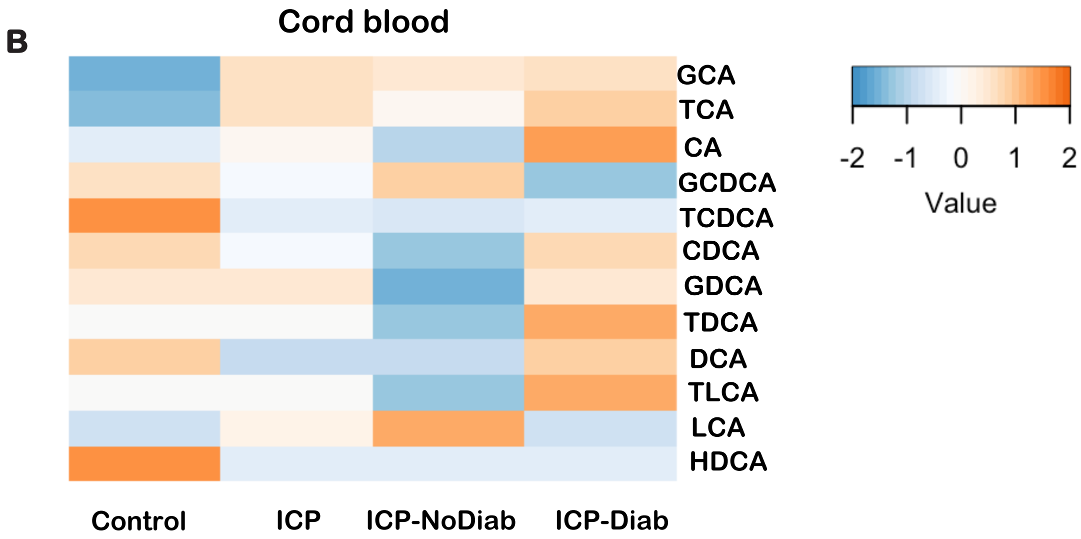


Supp Figure 1

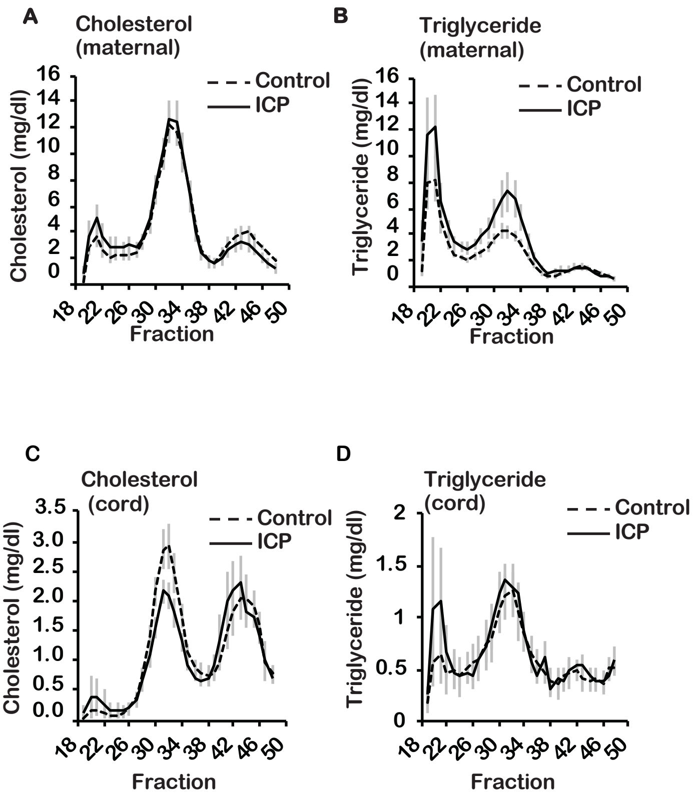


**Supplementary Figure 2**
