## Supplemental Table1-3 for "Unresolved alterations in bile acid composition and dyslipidemia in maternal and cord blood after ursodeoxycholic acid treatment for intrahepatic cholestasis of pregnancy"

**Table S1.** Maternal plasma BAs.

|  | Median [IQR] (μmol/l) |  |  |  |  | P value |  |  |  |
| --- | --- | --- | --- | --- | --- | --- | --- | --- | --- |
|  | **controls** | **ICP-all** | **ICP-NoDiab** | **ICP-Diab** |  | **ICP-all vs. controls** | **ICP-NoDiab vs. controls** | **ICP-Diab vs. controls** | **ICP-Diab vs. ICP-NoDiab** |
| UDCA | 0.003 [0.049] | 0.494 [1.111] | 0.135 [0.751] | 1.088 [1.371] |  | 0.059 | 0.183 | **0.008**** | 0.075 |
| TCDCA | 0.47 [0.96] | 0.61 [0.70] | 0.83 [0.72] | 0.41 [0.52] |  | 0.711 | 0.330 | 0.389 | 0.439 |
| GCDCA | 0.92 [0.51] | 0.83 [1.51] | 0.58 [2.40] | 1.06 [0.57] |  | 0.773 | 0.488 | 0.272 | 0.269 |
| CDCA | 0.04 [0.08] | 0.02 [0.04] | 0.01 [0.02] | 0.04 [0.04] |  | 0.091 | **0.018*** | 0.196 | 0.114 |
| TCA | 0.22 [1.42] | 0.69 [3.02] | 1.27 [2.13] | 0.46 [3.41] |  | 0.142 | 0.072 | 0.127 | 0.379 |
| GCA | 0.69 [0.91] | 1.26 [2.66] | 1.68 [1.87] | 1.26 [3.50] |  | 0.142 | 0.088 | 0.106 | 0.459 |
| CA | 0.05 [0.25] | 0.02 [0.05] | 0.02 [0.02] | 0.03 [0.15] |  | 0.196 | **0.036*** | 0.303 | 0.109 |
| TDCA | 0.69 [0.76] | 0.28 [0.37] | 0.28 [0.20] | 0.38 [0.71] |  | 0.536 | 0.185 | 0.398 | 0.269 |
| GDCA | 0.89 [1.10] | 0.75 [0.79] | 0.45 [0.60] | 1.00 [0.73] |  | 0.299 | **0.049*** | 0.413 | 0.083 |
| DCA | 0.50 [0.65] | 0.08 [0.10] | 0.07 [0.08] | 0.10 [0.14] |  | **0.002**** | **0.002**** | **0.017*** | 0.221 |
| TLCA | 0.006 [0.017] | 0.021 [0.024] | 0.015 [0.038] | 0.021 [0.013] |  | 0.196 | 0.173 | 0.086 | 0.341 |
| LCA | 0.001 [0.005] | 0.001 [0.007] | 0.007 [0.007] | 0.001 [0.000] |  | 0.696 | 0.136 | 0.364 | 0.082 |
| HDCA | 0.04 [0.09] | 0.00 [0.01] | 0.01 [0.03] | 0.00 [0.00] |  | **0.049*** | - | - | - |
| Total BAs (not including UDCA) | 5.19 [3.88] | 5.33 [6.46] | 5.83 [5.61] | 5.33 [6.14] |  | 0.837 | 0.446 | 0.383 | 0.439 |
| Total BAs (including UDCA) | 5.32 [3.84] | 6.90 [6.69] | 5.83 [5.55] | 7.45 [4.99] |  | 0.432 | 0.343 | 0.149 | 0.269 |
| Total CAs | 1.11 [2.80] | 2.35 [5.32] | 3.24 [3.34] | 1.71 [6.86] |  | 0.196 | 0.086 | 0.173 | 0.341 |
| Total CDCAs | 1.52 [1.29] | 1.61 [1.51] | 1.39 [3.14] | 1.62 [0.46] |  | 0.967 | 0.455 | 0.398 | 0.360 |
| Total DCAs | 2.27 [1.34] | 1.11 [0.99] | 0.90 [0.87] | 1.47 [1.70] |  | 0.100 | **0.021*** | 0.195 | 0.130 |
| CA/CDCA ratio | 0.73 [0.76] | 2.18 [1.52] | 2.36 [0.52] | 1.17 [1.11] |  | **0.017*** | **0.006**** | 0.061 | 0.178 |
| TBA/GBA ratio | 0.58 [0.24] | 0.60 [0.57] | 0.50 [0.72] | 0.64 [0.44] |  | 0.711 | 0.330 | 0.389 | 0.439 |
| CA/DCA ratio | 0.72 [1.89] | 2.58 [9.25] | 3.81 [13.08] | 1.24 [4.93] |  | 0.056 | **0.013*** | 0.135 | 0.141 |

*P<0.05, **P<0.01

We could not calculate Kruskal-Wallis for maternal HDCA, as it was below the limit of detection in all ICP-Diab patients.

**Table S2.** Cord plasma BAs.

|  | Median [IQR] (μmol/l) |  |  |  |  | P value |  |  |  |
| --- | --- | --- | --- | --- | --- | --- | --- | --- | --- |
|  | **controls** | **ICP-all** | **ICP-NoDiab** | **ICP-Diab** |  | **ICP-all vs. controls** | **ICP-NoDiab vs. controls** | **ICP-Diab vs. controls** | **ICP-Diab vs. ICP-NoDiab** |
| UDCA | 0.003 [0.018] | 0.047 [0.213] | 0.066 [0.336] | 0.047 [0.124] |  | **0.019*** | **0.014*** | **0.030*** | 0.378 |
| TCDCA | 1.40 [2.40] | 1.18 [1.61] | 2.02 [2.24] | 0.58 [0.57] |  | 0.583 | 0.279 | 0.055 | **0.018*** |
| GCDCA | 0.96 [1.30] | 1.50 [1.64] | 2.21 [1.78] | 0.58 [0.63] |  | 0.711 | 0.054 | 0.187 | **0.008**** |
| CDCA | 0.01 [0.01] | 0.02 [0.02] | 0.02 [0.02] | 0.02 [0.01] |  | 0.421 | 0.203 | 0.269 | 0.419 |
| TCA | 0.83 [0.73] | 1.21 [1.54] | 2.01 [4.31] | 1.01 [1.18] |  | 0.167 | **0.042*** | 0.235 | 0.165 |
| GCA | 0.32 [0.41] | 2.13 [1.98] | 2.62 [1.62] | 1.30 [2.15] |  | **0.013*** | **0.002**** | 0.084 | 0.083 |
| CA | 0.02 [0.03] | 0.06 [0.05] | 0.07 [0.05] | 0.06 [0.03] |  | **0.028*** | **0.021*** | **0.043*** | 0.379 |
| TDCA | 0.001 [0.000] | 0.001 [0.02] | 0.001 [0.010] | 0.001 [0.037] |  | 0.354 | 0.211 | 0.191 | 0.474 |
| GDCA | 0.01 [0.02] | 0.02 [0.04] | 0.01 [0.03] | 0.02 [0.06] |  | 0.641 | 0.460 | 0.166 | 0.151 |
| DCA | 0.02 [0.01] | 0.01 [0.01] | 0.01 [0.01] | 0.01 [0.01] |  | 0.299 | 0.130 | 0.226 | 0.360 |
| TLCA | 0.003 [0.005] | 0.004 [0.004] | 0.004 [0.002] | 0.003 [0.007] |  | 0.371 | 0.114 | 0.345 | 0.219 |
| LCA | 0.001 [0.000] | 0.001 [0.000] | 0.001 [0.004] | 0.001 [0.000] |  | 0.303 | - | - | - |
| HDCA | 0.003 [0.000] | 0.003 [0.002] | 0.004 [0.005] | 0.003 [0.000] |  | 0.549 | - | - | - |
| Total BAs (not including UDCA) | 5.36 [4.90] | 7.38 [5.90] | 9.33 [6.19] | 3.78 [4.67] |  | 0.340 | **0.030*** | 0.443 | **0.026*** |
| Total BAs (including UDCA) | 5.36 [4.89] | 7.61 [6.93] | 10.10 [5.55] | 4.00 [4.56] |  | 0.227 | **0.015*** | 0.497 | **0.018*** |
| Total CAs | 1.36 [1.49] | 3.66 [3.76] | 4.77 [5.79] | 2.58 [3.04] |  | **0.045*** | **0.009**** | 0.137 | 0.109 |
| Total CDCAs | 3.36 [3.20] | 3.38 [3.49] | 4.86 [1.33] | 1.18 [1.86] |  | 1 | 0.092 | 0.092 | 0.005** |
| Total DCAs | 0.02 [0.04] | 0.04 [0.06] | 0.04 [0.03] | 0.04 [0.10] |  | 1 | 0.416 | 0.416 | 0.341 |
| CA/CDCA ratio | 0.40 [0.44] | 1.31 [0.75] | 1.28 [0.94] | 1.31 [0.53] |  | **0.003**** | **0.016*** | **0.003**** | 0.269 |
| TBA/GBA ratio | 1.71 [0.66] | 0.88 [0.53] | 0.68 [0.48] | 1.02 [0.36] |  | 0.100 | **0.044*** | 0.119 | 0.304 |

*P<0.05, **P<0.01

We could not calculate Kruskal-Wallis for fetal LCA, as it was below the limit of detection in all controls and ICP-Diab, or fetal HDCA, as it was below the limit of detection in all ICP-Diab.

**Table S3.** Data collected from patient #21, an ICP-NoDiab patient who had previously undergone Roux-en-Y gastric bypass surgery. For ease of comparison, we also listed here the data from other ICP-NoDiab patients and controls, which are the same data reported throughout this manuscript. Glycemic parameters were not available for Patient #21.

|  | Control | ICP-NoDiab | Patient #21 |
| --- | --- | --- | --- |
| N | 7 | 6 | 1 |
| Maternal BAs (μmol/l) |  |  |  |
| UDCA | 0.003 [0.049] | 0.135 [0.751] | 0.386 |
| TCDCA | 0.47 [0.96] | 0.83 [0.72] | 0.09 |
| GCDCA | 0.92 [0.51] | 0.58 [2.40] | 1.80 |
| CDCA | 0.04 [0.08] | 0.01 [0.02] | 6.56 |
| TCA | 0.22 [1.42] | 1.27 [2.13] | 0.04 |
| GCA | 0.69 [0.91] | 1.68 [1.87] | 0.69 |
| CA | 0.05 [0.25] | 0.02 [0.02] | 4.20 |
| TDCA | 0.69 [0.76] | 0.28 [0.20] | 0.04 |
| GDCA | 0.89 [1.10] | 0.45 [0.60] | 0.69 |
| DCA | 0.50 [0.65] | 0.07 [0.08] | 3.39 |
| TLCA | 0.006 [0.017] | 0.015 [0.038] | 0.001 |
| LCA | 0.001 [0.005] | 0.007 [0.007] | 0.009 |
| HDCA | 0.04 [0.09] | 0.01 [0.03] | 0.15 |
| Total BAs (not including UDCA) | 5.19 [3.88] | 5.83 [5.61] | 17.66 |
| Total BAs (including UDCA) | 5.32 [3.84] | 5.83 [5.55] | 18.05 |
| Total CAs | 1.11 [2.80] | 3.24 [3.34] | 4.93 |
| Total CDCAs | 1.52 [1.29] | 1.39 [3.14] | 8.46 |
| Total DCAs | 2.27 [1.34] | 0.90 [0.87] | 4.12 |
| CA/CDCA ratio | 0.73 [0.76] | 2.36 [0.52] | 0.58 |
| TBA/GBA ratio | 0.58 [0.24] | 0.50 [0.72] | 0.05 |
| Cord BAs (μmol/l) |  |  |  |
| UDCA | 0.003 [0.018] | 0.066 [0.336] | 0.880 |
| TCDCA | 1.40 [2.40] | 2.02 [2.24] | 2.30 |
| GCDCA | 0.96 [1.30] | 2.21 [1.78] | 0.57 |
| CDCA | 0.01 [0.01] | 0.02 [0.02] | 0.36 |
| TCA | 0.83 [0.73] | 2.01 [4.31] | 0.37 |
| GCA | 0.32 [0.41] | 2.62 [1.62] | 0.12 |
| CA | 0.02 [0.03] | 0.07 [0.05] | 0.07 |
| TDCA | 0.001 [0.000] | 0.001 [0.010] | 0.063 |
| GDCA | 0.01 [0.02] | 0.01 [0.03] | 0.01 |
| DCA | 0.02 [0.01] | 0.01 [0.01] | 0.21 |
| TLCA | 0.003 [0.005] | 0.004 [0.002] | 0.001 |
| LCA | 0.001 [0.000] | 0.001 [0.004] | 0.001 |
| HDCA | 0.003 [0.000] | 0.004 [0.005] | 0.014 |
| Total BAs (not including UDCA) | 5.36 [4.90] | 9.33 [6.19] | 4.09 |
| Total BAs (including UDCA) | 5.36 [4.89] | 10.10 [5.55] |  |
| Total CAs | 1.36 [1.49] | 4.77 [5.79] | 0.55 |
| Total CDCAs | 3.36 [3.20] | 4.86 [1.33] | 3.24 |
| Total DCAs | 0.02 [0.04] | 0.04 [0.03] | 0.28 |
| CA/CDCA ratio | 0.40 [0.44] | 1.28 [0.94] | 0.17 |
| TBA/GBA ratio | 1.71 [0.66] | 0.68 [0.48] | 3.88 |
| Maternal lipid metabolism |  |  |  |
| Cholesterol (mg/dl) | 197.6 ± 16.1 | 197.5 ± 11.7 | 216.6 |
| HDL-cholesterol (mg/dl) by kit | 51.8 ± 8.8 | 34.4 ± 6.1 | 72.9 |
| Triglycerides (mg/dl) | 94.2 ± 9.3 | 95.1 ± 8.7 | 81.1 |
| Free fatty acids (mEq/l) | 0.44 ± 0.09 | 0.67 ± 0.11 | 0.49 |
| β-hydroxybutyrate (mmol/l) | 0.25 ± 0.04 | 0.56 ± 0.18 | 0.12 |
| Cord lipid metabolism |  |  |  |
| Cholesterol (mg/dl) | 95.9 ± 7.8 | 102.7 ± 9.8 | 101.1 |
| HDL-cholesterol (mg/dl) by kit | 24.2 ± 4.3 | 22.7 ± 2.2 | 23.5 |
| Triglycerides (mg/dl) | 8.1 ± 0.7 | 12.2 ± 1.4 | 9.8 |
| Free fatty acids (mEq/l) | 0.06 ± 0.01 | 0.13 ± 0.01 | 0.08 |
| β-hydroxybutyrate (mmol/l) | 0.16 ± 0.04 | 0.32 ± 0.10 | 0.06 |

**Supplementary Figure Legends**

**Supplementary Figure 1: Relative BA concentrations.** (A-B) Heat maps demonstrating relative concentrations of individual BAs, illustrated as z-score across groups in (A) maternal and (B) cord plasma from control and ICP subjects. To accompany the heat maps, absolute values, median, and interquartile range are provided in Tables S1-S2.

**Supplementary Figure 2.**  **Lipids in FPLC-fractionated maternal and cord blood plasma.** (A-B) Fractionation by FPLC of maternal blood plasma (n=5 controls and n=6 ICP), (C-D) Fractionation by FPLC of cord blood plasma (n=4 controls and n=5 ICP). Data show mean ± SEM.
